# Testicular microvascular flow is altered in Klinefelter syndrome and predicts circulating testosterone

**DOI:** 10.1101/2021.04.12.21255351

**Authors:** Francesco Carlomagno, Carlotta Pozza, Marta Tenuta, Riccardo Pofi, Luigi Tarani, Franz Sesti, Marianna Minnetti, Daniele Gianfrilli, Andrea M. Isidori

## Abstract

**Context:** Experimental studies on Klinefelter syndrome (KS) reported increased intratesticular testosterone (T) levels coexisting with reduced circulating levels. Abnormalities in testicular microcirculation have been claimed; however, no studies investigated *in vivo* testicular blood flow dynamics in humans with KS.

**Objective:** To analyze the testicular microcirculation in KS by contrast-enhanced ultrasonography (CEUS) and correlate vascular parameters with endocrine function.

**Design and Setting:** Prospective study. University Settings.

**Patients:** 51 testicular scans, 17 testes from 10 T-naïve subjects with KS and 34 testes from age-matched eugonadal men (CNT) who underwent CEUS for incidental nonpalpable testicular lesions.

**Main Outcomes:** CEUS kinetic parameters.

**Results:** CEUS revealed slower testicular perfusion kinetics in subjects with KS than in age-matched CNT. Specifically, the wash-in time (T_in_, *p* = 0.008), mean transit time (MTT, *p* = 0.008), time to peak (TTP, *p* < 0.001), and washout time (T_out 50%_, *p* = 0.008) were all prolonged. Faster testicular blood flow was associated with higher total T levels. Principal component analysis and multiple linear regression analyses confirmed the findings, and supported a role for reduced venous blood flow as independent predictor of total T levels.

**Conclusions:** Testicular venous blood flow is altered in KS and independently predicts T peripheral release.

## INTRODUCTION

Cardiovascular disease represents a major factor accounting for increased morbidity and mortality in subjects with Klinefelter syndrome (KS) (1,2). A Danish population study documented an approximately 2-fold increased risk of hospitalization because of circulatory diseases, which were present even before the diagnosis of KS, and a 4-fold increased risk of congenital heart malformations (2). In contrast, a British population study documented a 30% increase in mortality owing to circulatory diseases and a 7-fold increase in mortality owing to cardiovascular congenital anomalies compared with the background population (1). Contributing factors with respect to the heart and vasculature have been confirmed.

In the hearts of subjects with KS, an increased prevalence of mitral valve prolapse has been reported by some earlier studies (3,4), alongside diastolic dysfunction (5,6), chronotropic incompetence (6), and a shortened QTc interval (7). Vascular anomalies intrinsic to KS are evidenced by the finding of reduced luminal diameter of the main arterial blood vessels (namely, brachial artery, common carotid and femoral arteries, and abdominal aorta) (8), which is not explained by testosterone (T) levels or gonadal status and is coherent with a supposed X chromosome gene dosage effect; the opposite is indeed true in Turner syndrome, in which a tendency toward dilation of major arteries is observed (9). Furthermore, venous thrombosis is especially common in KS, with a 4-to 8-fold increased risk, and significantly contributes to the overall morbidity and mortality, independently of testosterone treatment and especially in young men (2,10,11).

Little is known regarding testicular vasculature and vascular abnormalities in subjects with KS and how these factors are possibly related to testicular function and frequent hypoandrogenism observed in this condition (12). Most intriguingly, a preliminary report from 2014 documented higher intratesticular T (ITT) levels in subjects with KS than those in normal controls, despite low serum T levels, resulting in a markedly increased ITT/T ratio (8 to 10 times higher in KS compared to controls), whereas the ITT/luteinizing hormone (LH) ratio was comparable among subjects with KS, patients with Sertoli-cell-only syndrome, and subjects with normal spermatogenesis (13). Under the assumption of normal Leydig cell function, the authors explored a “vascular” explanation for the lack of T release in the bloodstream and found evidence of vascular abnormalities in testis biopsies from adult male 41,XX^Y^ * vs 40,XY* mice. They observed a decreased blood vessel/testis surface ratio and, thus, speculated whether T production is not actually impaired in men with KS, but its release in the systemic circulation is hampered. They suggest that a reduced vascular bed is responsible for a lower T release into the bloodstream (13). The same group subsequently investigated the timing of vascular alterations during testicular development in mice, and found that such alteration are progressively acquired, with a lower number of small- and mid-sized blood vessels only in adult KS mice. They further conducted a study on testicular blood flow via contrast-enhanced ultrasonography (CEUS) and demonstrated reduced perfusion parameters, in favor of impaired testicular vascularization in KS mice (14).

Conversely, in human testicular biopsies obtained during testicular sperm extraction (TESE) procedure from prepubertal, pubertal, and adult subjects with KS, no differences were observed in blood vessel number or density when compared with age-matched control samples, except an increase in smaller vessels in prepubertal children only (15).

Color-flow systems used for the detection of vascular flow are challenged by the weak backscatter of blood relative to the tissue (−40 to −60 dB). The practical consequence of this challenge is limited sensitivity to low-velocity flow in smaller vessels (diameter < 2 mm), blood flow in deep vessels (diameter > 10 cm), and flow in regions with tissue motion (16). In the last decades, researchers have developed methods to detect perfusion using microbubbles to improve vascular flow on images. Following a bolus injection, the larger feeding vessels of the vascular tree can be visualized until the microcirculation fills. The use of intravascular CEUS has developed considerably and proved to be useful in many diagnostics fields (17-21). CEUS combines US with intravenous administration of ultrasonographic contrast medium comprising extremely small-sized US-detectable microbubbles (<10 μm), thus allowing visualization of the normal parenchymal microcirculation.

Based on these, the present study aimed to assess *in vivo* testicular blood flow using CEUS examination in adolescent and adult subjects with KS and to correlate vascular kinetics with endocrine function.

## MATERIALS AND METHODS

### Study participants

The PROMETEO study (PRecisiOn MEdicine to Target frailty of Endocrine-metabolic Origin) is a large multicenter prospective observational trial aimed at stratification risk and clinical score for the early recognition of endocrine frailty due to gonadal and adrenal disorders, focusing on cardiovascular (CV) and metabolic complications (Ethic Committee approval - RIF.CE 5563 prot. Prot n 727/19 of 12/09/2019). All participating patients provided written informed consent to the study.

Patients with KS or testicular lesions are at risk of developing hypogonadism, and, according to the protocol, they are screened with full hormonal assessment, plain testicular ultrasonography and, in case of altered echotexture, Contrast-Enhanced-Ultrasonography (CEUS). The inclusion criteria were as follows: 1) confirmed diagnosis of classic, non-mosaic 47, XXY KS based on karyotype analysis of leukocytes from peripheral blood; 2) availability of CEUS data; and 3) availability of concurrent clinical and hormonal data for correlation analyses. The exclusion criteria were as follows: 1) presence of other known genetic or chromosomal disorders; 2) anamnesis of cryptorchidism, testicular surgery, chemotherapy, or radiotherapy involving the pituitary gland or testes; and 3) current or previous T replacement therapy (TRT). Between, August 2020 and February 2021, 17 testicular scans, from 10 patients with KS and 34 testes from 34 eugonadal men who underwent CEUS for incidental nonpalpable testicular lesions being considered for analyses.

Control male subjects (CNT) were randomly matched to cases at a ratio of 2:1 testes by age (±1 year) and selected from subjects attending our outpatient clinic. They presented no clinical or biochemical evidence of hypogonadism, met the same inclusion and exclusion criteria described above (except KS diagnosis), and underwent testicular CEUS examination for the differential diagnosis of nonpalpable testicular lesions incidentally encountered during screening procedures conducted for varicocele and scrotal pain and during andrological prevention campaigns for high school and university students. All lesions were benign in nature at subsequent histology (n = 26 Leydig cell tumors, three epidermoid cysts, three focal fibrosis, one adenomatoid tumor, and one simple cyst).

### Hormonal evaluation

Blood samples were obtained via antecubital venous puncture in the early morning (07:30– 09:30 AM) after overnight fasting. Samples were centrifuged, and the serum was immediately frozen at −20°C. LH, follicle-stimulating hormone (FSH), sex hormone binding globulin (SHBG), and T levels were measured in duplicate using the chemiluminescent microparticle immunoassay (Architect System, Abbott Laboratories, IL, USA) with limits of detection of 0.07 mUI/mL, 0.05 mUI/mL, 0.28 nmol/L, and 0.1 nmol/L, respectively. The intra- and inter-assay coefficients of variation were as follows: 3.8% and 5.5% at 4.1 mUI/mL (LH), 3.6% and 5.4% at 3.2 mUI/mL (FSH), 5.65% and 9.54% at 8.8 nmol/L (SHBG), and 2.1% and 3.6% at 10.08 nmol/L (T), respectively. Reference ranges at our laboratory are as follows: LH < 1 (prepubertal) and 1.8–8.16 mUI/mL (adults); FSH < 1 (prepubertal) and 1.39–0.58 mUI/mL (adults); T < 3.2 (prepubertal) and 10.4–38.2 nmol/L (adults); and SHBG of 11.2–79.1 nmol/L. We employed Vermeulen’s formula to calculate the free T concentrations (cfT) from T and SHBG levels using a fixed albumin level of 4.3 g/dL (22) and derived the T/LH and cfT/LH ratios.

### CEUS

US examinations were performed using an iU22 unit (Philips, Bothell, Wash) with compound spatial and pulse inversion imaging and a 7–15-MHz wide-band linear probe. Axial and transverse scans of the testis were obtained based on scanning protocols used by Dogra et al (23) and Oyen (24). Representative images and loops were stored in the picture archiving and communication system for subsequent analysis. Testicular US was performed by two experienced sonographers with expertise in urogenital US and qualified in the use of CEUS (A.M.I. and C.P., both with >15 years of experience). All CEUS scans were obtained using the same machine with a low mechanical index (mechanical index = 0.07). Patients received intravenous injection of a 2.4-mL bolus (5 mg/mL) containing contrast material (SonoVue; Bracco, Milan, Italy), with a very rapid infusion, immediately followed by 10 mL of saline. Perfusion analyses were conducted using the quantification software QLAB (Philips). Curves were obtained by manually placing a region of interest (ROI) to entirely cover the testis, carefully excluding the lesion in the control group. ROIs did not change during the entire clip. Quantitative perfusion parameters were obtained using the gamma variate fit specifically designed for the analysis of perfusion imaging using microbubble agents; the function describes the overall transition of intensity value change during a contrast examination, starting with dark images, shortly followed by a steep increase in the early phase (wash-in) and enhancement with slow decrease over time at the end of the curve. The analyzed kinetic parameters included wash-in time (T_in_, in seconds), coefficient of the wash-in slope (Slope_in_, in dB/second), peak intensity (PI, in dB), time to peak (TTP, in seconds), mean transit time (MTT, in seconds), time of washout 50% (T_out 50%_, in seconds), and area under the curve (AUC, in arbitrary units) **(Figure 1)**. The Microvascular Imaging (MVI) plug-in for measuring changes in the image from frame to frame, suppressing background tissue signals, and capturing additional contrast data was employed to dramatically enhance vessel conspicuity and map contrast agent progression.

**Figure 1.**
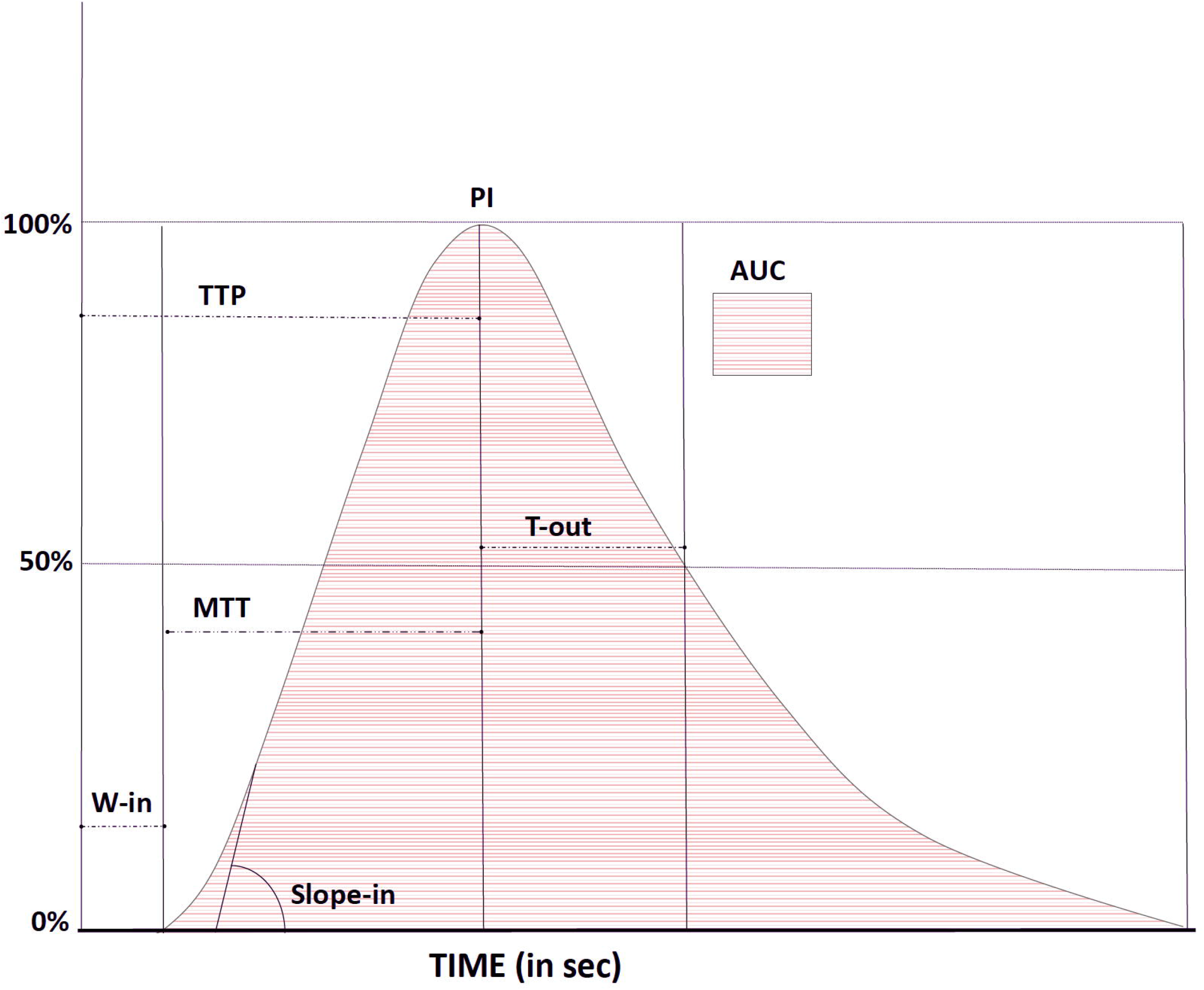
Graphic representation of the time/intensity curve and calculated perfusion parameters: *wash-in time (W-in)*, the time when the testis first started to enhance, measured in seconds; *time to peak (TTP)*, the time needed to reach the peak intensity, measured in seconds; *mean transit time (MTT)*, the time difference between the time needed to reach the peak intensity and the time of the start of the enhancement for the ROI, measured in seconds; *peak intensity (PI)*, the maximum enhancement of the ROI, measured in decibel (dB); washout *time (T-out)*, the time difference between the respective 50% peak intensity values in the washout and peak intensity value, measured in seconds; *area under the curve (AUC)*, intensities of the whole period of enhancement, measured in arbitrary units; S*lope in*, the coefficient of the wash-in slope and reflects the mean blood flow velocity in the region of interest, measured in in dB/second.

### Statistical analysis

Data were expressed as means (M) and standard deviations or medians (m) and 25%–75% interquartile ranges (IQRs), as appropriate. Data distribution was visually inspected by analyzing the respective histograms and normality plots. The normally distributed data were analyzed with independent samples *t*-tests. The non-normally distributed data were analyzed using Mann–Whitney *U* tests and linear regression, using bootstrapping for 2000 samples and bias corrected accelerated (BCa) 95% confidence intervals. Spearman’s correlation coefficient (r_S_) was used for correlation analyses of non-normally distributed data. The level of statistical significance was set at a *P-*value of < 0.05. Data were visually represented with box-whisker plots as m (black lines), 25%–75% IQR (boxes), and 2.5–97.5th percentiles (whiskers). A principal component analysis (PCA) was used to reveal the internal structure of CEUS kinetic parameters (T_in_, Slope_in_, PI, TTP, MTT, T_out 50%_ and AUC) through the impartial extration of a number of uncorrelated parameters, called principal components (PCs), to retain most of the information present in the original data. All statistical computations were conducted with the IBM SPSS Statistics for Windows (version 25, IBM Corp.) and GraphPad Prism for Windows (version 8.3.0, GraphPad Software, LLC).

## RESULTS

Age, testicular volumes, endocrine function, and CEUS parameters of patients with KS and age-matched control subjects are presented in **Table 1**.

**Table 1.** General, hormonal, and CEUS data of the testes of subjects with KS and age-matched healthy controls (m [25%–75% IQR])

|  | KS subjects | Healthy controls | *P* |
| --- | --- | --- | --- |
| n | 10 | 34 |  |
| Testes | 17 | 34 |  |
| Age, years* | 31.7 ± 12.04 | 31.6 ± 6.29 | 0.82 |
| Testicular volume, mL# | 3.5 (2.9–3.7) | 17.1 (14.4–21.2) | **<0.001** |
| **Endocrine function** |  |  |  |
| LH, mUI/mL | 12.03 (10.58–16.46) | 2.9 (2.25–3.69) | **<0.001** |
| FSH, mUI/mL | 29.12 (22.66–38.53) | 3.15 (1.64–5.69) | **<0.001** |
| T, nmol/L | 13.24 (10.92–18.2) | 21.47 (17.49–25.36) | **0.02** |
| SHBG, nmol/L | 35.4 (30.85–42.25) | 42.3 (29.95–56.1) | 0.29 |
| cfT, nmol/L | 0.35 (0.16–0.42) | 0.41 (0.34–0.47) | 0.154 |
| T/LH ratio | 1.14 (0.59–1.5) | 7.01 (4.94–10.2) | **<0.001** |
| cfT/LH ratio | 0.03 (0.01–0.04) | 0.15 (0.1–0.2) | **<0.001** |
| **CEUS parameters** |  |  |  |
| T<sub>in</sub>, seconds | 9.36 (7.54–12.73) | 7.42 (5.45–8.82) | **0.008** |
| Slope<sub>in</sub> | 0.25 (0.17–0.4) | 0.28 (0.22–0.42) | 0.441 |
| PI, dB | 2.86 (2.02–3.65) | 2.41 (1.77–3.22) | 0.184 |
| MTT, seconds | 11.8 (10.66–17.53) | 10.46 (8.21–12.75) | **0.008** |
| TTP, seconds | 42.3 (37.85–50.47) | 35 (26.8–38.54) | **<0.001** |
| T<sub>out 50%</sub>, seconds | 31.51 (21.93–38.76) | 23.41 (16.92–29.95) | **0.008** |
| AUC | 116.85 (94.26–158.71) | 94.48 (58.41–156.59) | 0.129 |

*Note:* Parameters marked by \* show a normal distribution, are presented as mean ± standard deviation, and have been tested via independent samples *t*-test. # Volume of testes that underwent CEUS examination are reported. Significant *P*-values are presented in bold.
Abbreviations: **KS**, Klinefelter syndrome; **CEUS**, contrast-enhanced ultrasonography, **T<sub>in</sub>**, wash-in time (in seconds); **Slope<sub>in</sub>**, coefficient of the wash-in slope (in dB/second); **PI**, peak intensity (in dB); **MTT**, mean transit time (in seconds); **TTP**, time to peak (in seconds); **T<sub>out 50%</sub>**, time difference between the PI value and 50% of the PI value in the wash-out (in seconds); **AUC**, area under the curve (in arbitrary units)

As expected, compared with the CNT group, subjects with KS presented significantly reduced testicular volumes (*p* < 0.001), higher gonadotropin levels (*p* < 0.001 for both LH and FSH), and reduced serum T levels (*p* = 0.02) as well as both T/LH and cfT/LH ratios (*p* < 0.001 for both) but similar SHBG and cfT levels. Specifically, of the 10 enrolled subjects with KS, four were hypoandrogenemic (T levels < 10.4 nmol/L), whereas the remaining six individuals presented T levels within the reference range.

### CEUS parameters of testicular parenchymal vascularization

The analysis of CEUS parameters showed slower testicular perfusion kinetics in subjects with KS than in the age-matched CNT group. Specifically, there was a significantly longer wash-in phase of contrast microbubbles and mean testicular transit time, as evidenced by slower T_in_ (*p* = 0.008), MTT (*p* = 0.008), and TTP (*p* < 0.001) intervals. The washout phase was also significantly slower, with a prolonged T_out 50%_ (*p* = 0.008). No significant differences were observed with regard to the Slope_in_, PI, or AUC. Vascular kinetics are presented in **Figure 2**, which depicts a representative examination in a patient with KS (upper panels) and CNT subject (lower panels), alongside perfusion curves (subject with KS in red and CNT subject in light blue).

**Figure 2.**
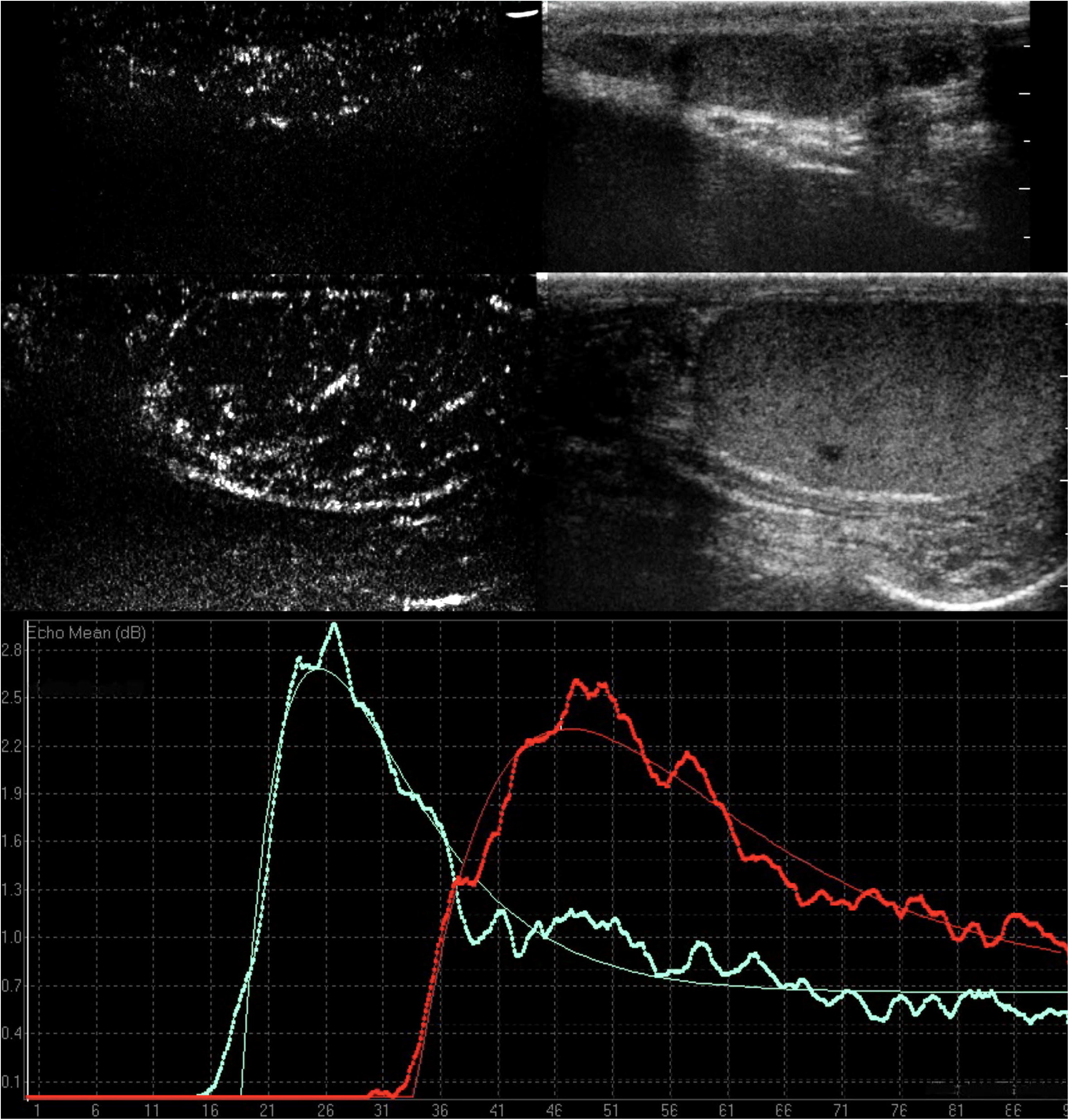
Quantitative contrast-enhanced US of a KS testis (upper panels) and a CNT testis (lower panels). Contrast pulse inversion harmonic time–intensity curves of the KS testis (in red) and the CNT testis (in light blue) show a slower T_in_ and TTP and longer MTT and T_out 50%_ in the KS testis than in the CNT testis.

MVI depiction of the differences between a KS testis (left) and CNT testis (right) is displayed in **Figure 3**: at 30 seconds (panel B), the vascular architecture of the CNT testis is already apparent with an organized and regular pattern, whereas the KS testis vessels are only visualized at 45 seconds (panel D), showing an irregular and chaotic vascular pattern.

**Figure 3.**
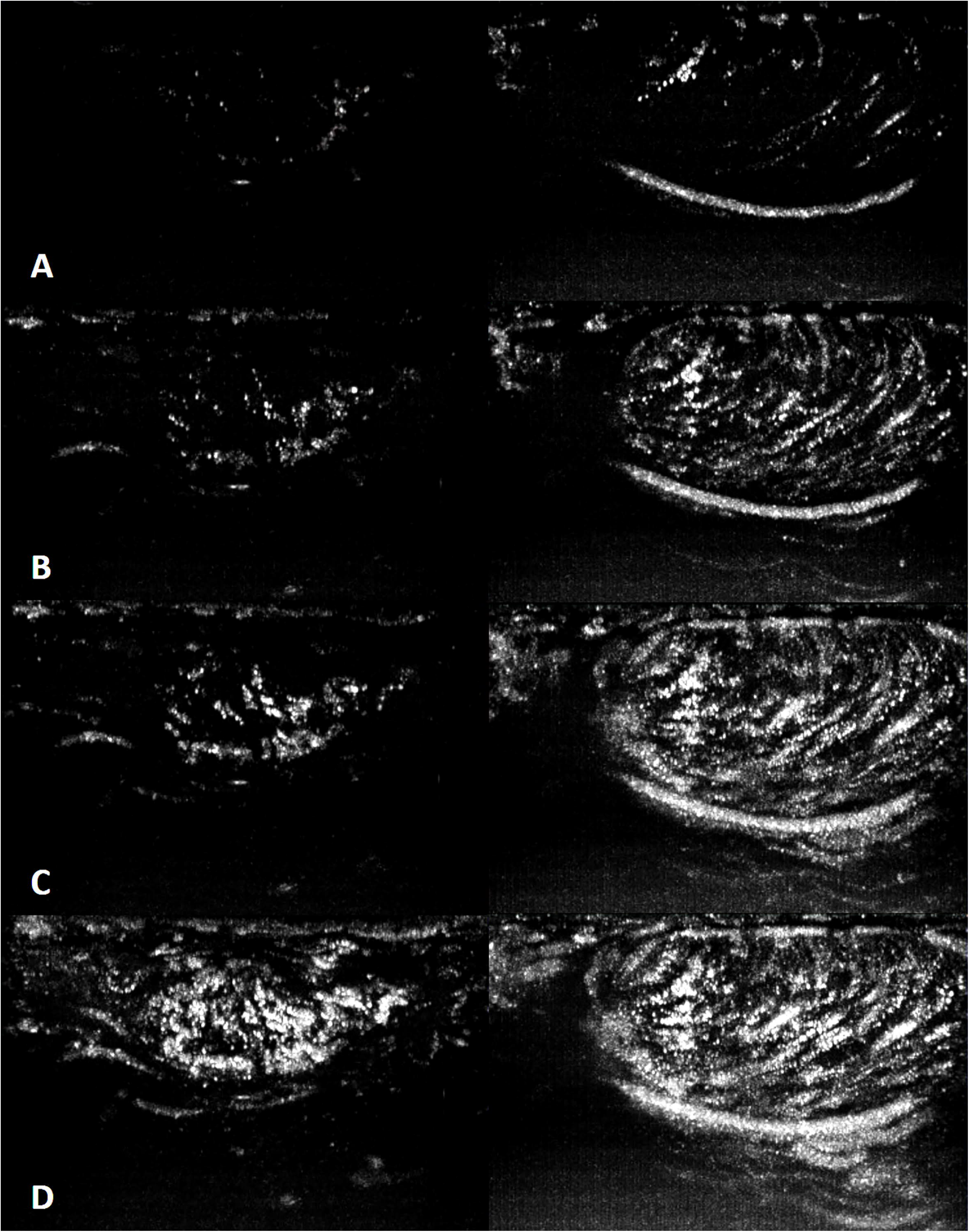
Microvascular Imaging (MVI) frames of KS testis (left) and CNT testis (right) at 20 (panel A), 30 (panel B), 45 (panel C) and 90 seconds (panel D) after contrast injection clearly show the different behavior of intratesticular vessels. At 30 seconds, the CNT testis is already filled with Sonovue, its vascular architecture is clearly identified, and the progression is organized and regular; conversely, KS testis vessels are visualized at 45 seconds, and at the end of the examination, the map shows irregular and chaotic vascularization.

### Correlation analyses of testicular vascularization and endocrine function

A correlation analysis among age, testicular volume (of the examined testis), and CEUS parameters showed no significant associations (data not shown), except for TTP, which showed a positive correlation with age (r_S_ = 0.31, *p* = 0.046) and testicular volume (r_S_ = −0.38, *p* = 0.003).

To examine the interrelationship between testicular vascular parameters assessed by CEUS and testicular endocrine function, we further conducted a correlation analysis in subjects with KS and CNT. Complete results are summarized in **Table 2**.

**Table 2.**
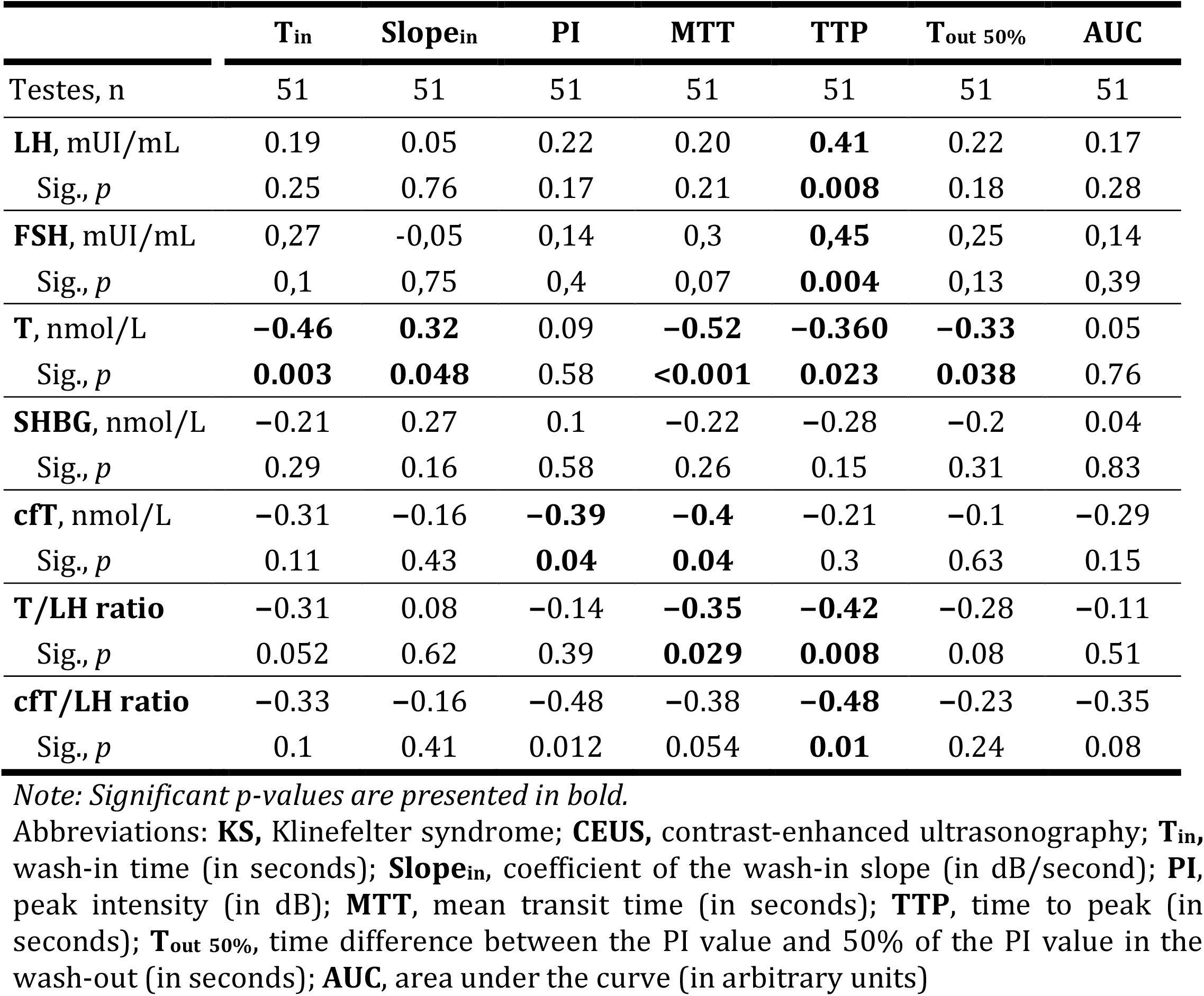
Spearman correlation coefficients between CEUS-derived testicular vascular parameters and endocrine function in subjects with KS and healthy control subjects

|  | **T<sub>in</sub>** | **Slope<sub>in</sub>** | **PI** | **MTT** | **TTP** | **T<sub>out</sub> 50%** | **AUC** |
| --- | --- | --- | --- | --- | --- | --- | --- |
| Testes, n | 51 | 51 | 51 | 51 | 51 | 51 | 51 |
| **LH**, mUI/mL | 0.19 | 0.05 | 0.22 | 0.20 | **0.41** | 0.22 | 0.17 |
| Sig., *p* | 0.25 | 0.76 | 0.17 | 0.21 | **0.008** | 0.18 | 0.28 |
| **FSH**, mUI/mL | 0.27 | -0.05 | 0.14 | 0.3 | **0.45** | 0.25 | 0.14 |
| Sig., *p* | 0.1 | 0.75 | 0.4 | 0.07 | **0.004** | 0.13 | 0.39 |
| **T**, nmol/L | **-0.46** | **0.32** | 0.09 | **-0.52** | **-0.360** | **-0.33** | 0.05 |
| Sig., *p* | **0.003** | **0.048** | 0.58 | **<0.001** | **0.023** | **0.038** | 0.76 |
| **SHBG**, nmol/L | -0.21 | 0.27 | 0.1 | -0.22 | -0.28 | -0.2 | 0.04 |
| Sig., *p* | 0.29 | 0.16 | 0.58 | 0.26 | 0.15 | 0.31 | 0.83 |
| **cfT**, nmol/L | -0.31 | -0.16 | **-0.39** | **-0.4** | -0.21 | -0.1 | -0.29 |
| Sig., *p* | 0.11 | 0.43 | **0.04** | **0.04** | 0.3 | 0.63 | 0.15 |
| **T/LH ratio** | -0.31 | 0.08 | -0.14 | **-0.35** | **-0.42** | -0.28 | -0.11 |
| Sig., *p* | 0.052 | 0.62 | 0.39 | **0.029** | **0.008** | 0.08 | 0.51 |
| **cfT/LH ratio** | -0.33 | -0.16 | -0.48 | -0.38 | **-0.48** | -0.23 | -0.35 |
| Sig., *p* | 0.1 | 0.41 | 0.012 | 0.054 | **0.01** | 0.24 | 0.08 |

*Note: Significant p-values are presented in bold.*
Abbreviations: **KS**, Klinefelter syndrome; **CEUS**, contrast-enhanced ultrasonography; **T<sub>in</sub>**, wash-in time (in seconds); **Slope<sub>in</sub>**, coefficient of the wash-in slope (in dB/second); **PI**, peak intensity (in dB); **MTT**, mean transit time (in seconds); **TTP**, time to peak (in seconds); **T<sub>out</sub> 50%**, time difference between the PI value and 50% of the PI value in the wash-out (in seconds); **AUC**, area under the curve (in arbitrary units)

Among the hormonal parameters considered, serum T levels showed the strongest relationship with vascular parameters, correlating positively with Slope_in_ (*p* = 0.048) and inversely with T_in_ (*p* = 0.003), MTT (*p* < 0.001), TTP (*p* = 0.023), and T_out_._50%_ (*p* = 0.038). These parameters (except Slope_in_) were deemed significantly different between the KS and CNT groups. Opposite significances were found for LH and FSH levels solely with the TTP. Serum cfT levels were inversely correlated with PI and MTT values, whereas the T/LH and fT/LH ratios were inversely correlated with TTP and the T/LH ratio with MTT).

### Principal component analysis of CEUS parameters

A PCA was conducted on CEUS kinetic parameters with oblique rotation (direct oblimin). The Kaiser-Meyer-Olkin measure verified the sampling adequacy for the analysis, KMO = 0.63, and all KMO values for individual items were greater 0.5. An initial analysis was run to obtain eigenvalues for each factor in the data. Two components had eigevalues over Kaiser’s criterion of 1 and in combination explained 88.6% of the variance. The scree plot showed inflexion that justified retaining two components. The first PC (PC1) comprised the following loadings: T_in_ (0.98), Slope_in_ (−0.516), TTP (0.793), MTT (0.952) and T_out 50%_ (0.793); this PC showed an eigenvalue of 3.76 and explained 53.8% of the total variance. The second PC (PC2) comprised the following loadings: Slope_in_ (0.881), PI (0.967) and AUC (0.830; this PC showed an eigenvalue of 2.44 and explained 34.8% of the total variance. The items that cluster on the same component suggest that PC1 represents time-dependent behaviour, while PC2 represents intensity-dependent behaviour.

### Multiple regression analysis of PCs and total T levels

A multiple linear regression analysis was conducted to determine whether vascular parameters were able to independently predict total T levels in KS and CNT subjects; complete results are reported in **Table 3**. Total T levels were predicted positively by bitesticular volume (*p* = 0.017) and negatively by PC1 (*p* = 0.04); age and PC2 were not found to be significant predictors.

**Table 3.** Linear model of predictors of total testosterone in KS subjects and healthy controls after bootstrapping.

|  | *b* | *SE* | *p* |
| --- | --- | --- | --- |
| **Total testosterone, nmol/L** |  |  |  |
| Constant | 12.257 (5.876, 20.237) | 4.139 | 0.006 |
| Age, years | 0.107 (-0.15, 0.346) | 0.118 | 0.36 |
| Bitesticular volume, mL | 0.174 (0.03, 0.31) | 0.067 | **0.017** |
| PC1 | -2.467 (-4.757, -0.1) | 1.186 | **0.04** |
| PC2 | 0.740 (-1.446, 3.378) | 1.267 | 0.58 |

*Note:* 95% bias corrected accelerated (Bca) confidence intervals from 2000 bootstrapped samples are reported in parentheses. Significant *P*-values are presented in bold.
Abbreviations: **KS** = Klinefelter syndrome; **PC** = principal component; **SE** = standard error.

## DISCUSSION

The presence of increased cardiovascular risk in KS is a well-established fact and has been documented in population studies, which have described an increase in morbidity and mortality (1,2). The contributing factors are less known, and although T probably plays a role as a vasodilator, with an apparent protective effect with regard to atherogenesis (25), the aneuploidy condition per se seems to be involved. This consideration occurs owing to the higher incidence of congenital heart malformations (2) and the finding of reduced luminal diameter of the main arteries in subjects with KS, independent of T levels or gonadal status (8). Whether these anomalies extend to testicular vasculature or whether testicular function is affected is a matter of debate.

Although a Doppler US study initially described high-resistance intratesticular blood flow in subjects with KS (26), significant findings were obtained in 2014 from a preliminary report describing surprisingly increased ITT levels in subjects with KS compared with healthy controls (almost 10 times higher) (13). This result prompted authors to search for a possible explanation to this phenomenon, discarding the idea of impaired Leydig cell function in KS. The histological finding of a low blood vessel/testis surface ratio in a mouse model of KS led to the hypothesis that T release, rather than its production, is actually impaired in KS, by means of a slower and reduced testicular blood flow. These results were confirmed in a subsequent study, in which a reduction in small and medium-sized blood vessels in adult 41,XX^Y^ * mice was described, and the CEUS study of those testes demonstrated reduced perfusion parameters, favoring impaired testicular vascularization in KS (14). However, these results were inconsistent with the findings of another histological study in human KS testicular biopsies from prepubertal, peripubertal, and adult subjects (15).

The present study examined for the first time *in vivo* testicular vascularization in human subjects with KS by means of CEUS compared with age-matched controls. Subjects with KS were characterized by significantly slower testicular perfusion kinetics, in both the wash-in and washout phases **(Figure 2)**, resulting in prolonged testicular MTT; the testicular Slope_in_ was not significantly different between the two groups, possibly because of the lack of differences in the number of larger arteries (14), in which microbubbles first appear; the PI and AUC were also not different between the two groups.

Strikingly, faster testicular vascularization times were associated with better endocrine testicular function, especially with serum T levels, which showed a strong relationship with vascular parameters, correlating positively with Slope_in_ and inversely with T_in_, MTT, TTP, and T_out_._50%_ as well as gonadotropins and T/LH and cfT/LH ratios. Using MVI analysis, we observed vessel conspicuity during contrast agent progression and noted the normal vascular architecture of CNT compared with subjects with KS, in whom small vessels were not clearly seen.

To assess the individual contribution of vascular flow on total T levels we first performed a PCA on CEUS parameters in KS and CNT subjects: two PCs were identified, namely PC1, representing time-dependent behaviour and mostly indicative of venous blood flow, and PC2, representing intensity-dependent behaviour, mainly attributable to large arteries blood flow (27). Of note, PC1 was constituted by the same CEUS parameters upon which KS subjects significantly differed from the CNT cohort. We subsequently proceeded to conduct a multiple linear regression analysis to verify the independent role of vascular blood flow on testicular endocrine function, which showed venous blood flow (PC1) and bitesticular volume to be independent predictors of total T levels.

To our knowledge, this is the first demonstration that altered intratesticular vasculature correlates with altered peripheral hormonal release. If the feedback loop is indeed intact, an inefficient hormone release in the circulation could be masked by an increase in LH, stretching the hypothalamic–pituitary–testicular axis to the maximum but still resulting in lower circulating T levels. Therefore, the higher ITT/T ratio observed in KS, according to the data published by Tuttelmann et al. (13), would not be a sign of Leydig cell failure but a sign of failure of the interaction between the T source and distribution system. PCA and linear regression analyses further point towards a role for reduced venous blood flow in regulating T release from the testes.

Our study has some limitations. First, the sample size and age distribution of enrolled men with KS are small, resulting in 17 testes from adolescents and adults; however, this is in line with the preliminary nature of our study. Second, as a control group, we included subjects undergoing CEUS because of nonpalpable lesions of < 0.5-cm diameter which were benign at definitive histological evaluation. Therefore, we do not expect testicular parenchymal vascularization to be affected in these men, although this possibility cannot be completely excluded.

Lastly, kinetic parameters depend on the manufacturer of the US unit, and although our software provides automatic time–intensity curves measurements, it is possible that analyses conducted using different software on other US systems provide different values.

Further studies are warranted to evaluate testicular KS vascularization in the prepubertal and peripubertal timing, establish whether vascular abnormalities are already present at an earlier stage, and determine whether vascular dynamics can be useful in predicting the “testicular catastrophe” occurring in KS and scheduling TESE.

In conclusion, our study provides the first demonstration of impaired testicular vascularization kinetics in men with KS. These alterations are possibly related to T release in the peripheral circulation.

## Data Availability

All data will be made available upon reasonable request.

https://isidorilab.com/home

